# COVID-19 vaccines and evidence-based medicine

**DOI:** 10.1101/2021.06.28.21259039

**Authors:** Andrew Larkin, Howard Waitzkin

## Abstract

**OBJECTIVE:** To clarify efficacy, effectiveness, and harm of available vaccines for COVID-19, using measures in evidence-based medicine (EBM) that, in addition to relative risk reduction, consider absolute risk reduction and variations in baseline risks.

**DESIGN:** Systematic review of studies that have considered impacts of vaccines in relation to baseline risks. Calculation of risk reduction and harms from published data in two random controlled trials and one population-based implementation study. Analysis of risk reductions in geographical areas with varying baseline risks. Comparison of results concerning COVID-19 vaccine and selected prior vaccines.

**SETTING:** Random controlled trials of Pfizer and Moderna vaccines in multiple countries; population-based study using Pfizer vaccine in Israel. Counties with varying baseline risks in the United States; states with varying baseline risks in India.

**PARTICIPANTS:** 43,448 and 30,420 subjects in the random controlled trials; 1,198,236 subjects in the population-based study.

**INTERVENTIONS:** Multi-site random controlled trials of vaccine efficacy; population-based administration of vaccine with determination of effectiveness by comparison of vaccinated and unvaccinated subjects.

**MAIN OUTCOME MEASURES:** Relative risk reduction (RRR), absolute risk reduction (ARR), number needed to be vaccinated to prevent one symptomatic infection (NNV), absolute risk of the intervention (ARI), and number needed to harm (NNH).

**RESULTS:** A systematic review of literature in medicine and public health showed very few reports regarding ARR, NNV, ARI, and NNH; use of these indicators to compare benefits versus harms; or analysis of these EBM indicators in the context of varying baseline risks. From data in the two random controlled trials and one population-based study, calculated ARR was approximately 1 percent (as compared to RRR of 50 to 95 percent), and NNV was in the range of 100 to 500. In comparisons of ARR and NNV versus ARI and NNH, benefits and harms were not markedly different. From a sensitivity analysis of ARR and NNV in population groups with varying baseline risks, the effectiveness of vaccines as measured by ARR and NNV was substantially higher in regions with high as compared to low baseline risks. The ARR for COVID-19 vaccines was somewhat smaller and the NNV somewhat larger than achieved by some vaccines to prevent influenza and smallpox.

**CONCLUSION:** The efficacy and effectiveness of major COVID-19 vaccines, as measured by RRR, are impressive. As measured by ARR and NNV, which take into account variation in baseline risks, the effectiveness of the vaccines is substantially higher in areas with higher baseline risks. This finding can contribute to educational efforts, informed consent procedures, and policy making about priorities for vaccine distribution, especially under conditions of access barriers linked to poverty and inequality.

**WHAT IS ALREADY KNOWN ON THIS TOPIC:** Major COVID-19 vaccines so far have shown impressive efficacy in random controlled trials and effectiveness in population-based studies. To determine efficacy and effectiveness, these studies have used relative risk reduction (RRR), which shows the difference in event rate between those receiving and not receiving a vaccine. Reports of efficacy and effectiveness have not yet clarified other key indicators from evidence-based medicine (EBM) that consider variations baseline risks. Such indicators include measures of benefits such as absolute risk reduction (ARR) and number needed to be vaccinated (NNV), as well as measures of harm such as absolute risk of the intervention (ARI) and number needed to harm (NNH).

**WHAT THIS STUDY ADDS:** For COVID-19 vaccines, calculated ARR is somewhat lower and NNV somewhat higher than for certain prior vaccines such as those for influenza and smallpox. Indicators of harm for COVID-19 vaccines, as measured by ARI and NNH, appear to be in the same order of magnitude as indicators of benefit such as ARR and NNV. The effectiveness of COVID-19 vaccines, as measured by ARR and NNV, is substantially higher in geographical areas with high baseline risk, compared to areas with low baseline risk.

These findings can assist in informed consent procedures, educational efforts, and priority setting in policies about distribution of vaccines, especially in the context of access barriers related to poverty and inequality.

## Introduction

Among its challenges, the COVID-19 pandemic has worried some practitioners, researchers, and teachers who have tried to promote evidence-based medicine (EBM). As general internists, we have written this article to help clarify the answers to some puzzling questions about the use of EBM approaches in understanding the benefits and possible harms of vaccines for COVID-19. We also are trying to develop clear ways to explain EBM approaches concerning the vaccines to patients, the public, and policy makers responsible for decision making about the distribution of the vaccines to populations.

For instance, through our generally supportive though sometimes critical study and teaching of EBM, we have learned and taught that absolute risk reduction (ARR) and the number needed to be treated or vaccinated (NNT, NNV), in addition to relative risk reduction (RRR), are fundamental measures in evaluating a new treatment or other clinical interventions such as a vaccine. In a classic statement of this principle, leaders of EBM argued^1^:

> In reviewing the different ways that benefit and harm can be expressed, we conclude that the absolute risk reduction is superior to the relative risk reduction because it incorporates both the base-line risk and the magnitude of the risk reduction. Its reciprocal, the number needed to be treated, expresses the absolute risk reduction in a manner that is easily understood by clinicians, and can be used to describe the harm as well as the benefits of therapy and other clinical maneuvers.

A widely used textbook on EBM, in its 2019 edition, similarly favors the reporting of ARR^2^: “Thus, the ARR is a more meaningful measure of treatment effects compared with the RRR.” A publication of the U.S. Food and Drug Administration (FDA) also recommends reporting both ARR and RRR in assessments of treatment or vaccine efficacy^3^:

> Provide absolute risks, not just relative risks. Patients are unduly influenced when risk information is presented using a relative risk approach; this can result in suboptimal decisions. Thus, an absolute risk format should be used.

Yet despite these recommendations, publications of the major COVID-19 vaccine evaluations, both random controlled trials (RCTs) and population-based assessments, have reported findings about RRR only. ARR and NNV have been reported and considered in guidelines concerning other vaccines such as influenza.^4,5,6^ But ARR and NNV have not been reported regularly or used to recommend policies regarding COVID-19 vaccines. (In the medical literature, the following terms are used interchangeably: “numbers needed to treat” and “numbers needed to be treated”; and “numbers needed to vaccinate” and “numbers needed to be vaccinated.” We consistently use NNV, referring to the number needed to be vaccinated in order to prevent one event, such as a symptomatic infection with COVID-19.) Moreover, to compare benefits and harms from an intervention, the number needed to harm one person (NNH) can be compared to the NNV, but this comparison also has not been reported for the major COVID-19 vaccines.

Because of these concerns, we have tried to address the following questions:

1. What are the RRRs and ARRs achieved by available vaccines, and what is the NNV in order to prevent one symptomatic infection?
2. To assess benefits versus harms, what is the NNH for the vaccines, compared to NNV?
3. How does the NNV vary in populations with different baseline risks of disease, and what are the implications for strategies of vaccine distribution?

Beyond trying answer these three questions, we aimed to compare measures of benefits and harms to those of prior vaccines generally considered successful. Finally, we hoped to prepare a simple explanation of our findings that would assist in educational efforts, informed consent procedures, and policy decisions about vaccine distribution.

By limiting our report to these questions and goals, we chose not to deal here with other troubling issues pertinent to EBM. These issues include but are not limited to the accuracy of test results (the operating characteristics of current tests in terms of sensitivity and specificity, as well as pre-test and post-test probabilities as informed by Bayes’ theorem); underestimates of disease and death due to incomplete case identification; the costs of vaccines in relation to public-sector financial support for private pharmaceutical corporations; the effects of racism, poverty, inequality, intellectual property rights, and other structural conditions that block access to vaccines and medical care; and the disorganized policies of some governments and health institutions.

## Methods

### Systematic review

During April 2021 we searched PubMed for articles combining COVID and SARS-CoV-2 separately with each of the following terms: ARR, RRR, NNT, NNV, risk-benefit, and NNH. As feasible, we applied the PRISMA 2020 guideline in planning and executing our search.^7^ Initially, because we anticipated that few publications would be revealed by this search, we did not introduce additional selection or exclusion criteria. Because we found so few publications containing these terms, we requested that experienced reference librarians at the University of New Mexico Health Sciences Library and Informatics Center do an independent search of the literature; they found no additional publications beyond those located through our initial search. We also supplemented this search of the medical and public health literature by using a two general search engines, DuckDuckGo and Google, with the same search terms. Through notes and a spreadsheet, we separately assessed each article to clarify answers, if any, to the 3 questions above. Then we communicated about our findings and resolved differences in interpretation through discussion.

### Assessment of vaccines

For our assessment of vaccine trials, we applied the definitions in Box 1, which were developed in EBM and in systematic reviews of vaccine evaluations.^1,2,8,9,10,11^

We used these EBM measures to analyze reports from three influential studies. Two of the studies, concerning the Pfizer^12^ and Moderna^13^ vaccines, were efficacy trials, showing the impact of an intervention in an RCT. An effectiveness trial, in Israel, was an observational study showing the “real world” impact of vaccine on population-level outcomes.^14^ Using published data from each study, we calculated RRR, ARR, NNV, ARI, and NNH. We then compared benefits and harms, using NNV and NNH.

To clarify implications for policies about vaccine distribution and accessibility, we performed a sensitivity analysis that considered ARR and NNV in regions with differing baseline risks of disease. We obtained the baseline risks for selected U.S. counties from the Johns Hopkins University dashboard and for selected states of India from the New York Times dashboard.^15,16^

To place COVID vaccines in an historical context, we compared available data to those achieved by previous vaccines. For these comparisons, we focused on benefits as measured by NNV, calculated from the above publications concerning vaccines for COVID-19 and vaccines for several other viral diseases.

### Patient and public involvement

We tried to illustrate how these findings and policy implications can be explained simply to patients, policy makers, and the general public. In designing the study, we took into account questions that we have received frequently from patients, colleagues, and members of our communities. Due to the rapid changes resulting from the COVID-19 pandemic, we chose not allocate time for more systematic sampling of these groups to clarify the frequencies of different concerns. We plan to disseminate the findings widely to the public and policy makers through press releases, media contacts, institutional websites, blogs, list serves, personal communications, and social media tools.

## Results

### Literature review

We found very few publications that referred to ARR and NNV in research on COVID-19 vaccines. In the medical literature, one article oriented to EBM criticized the deficit in reporting of these measures in the major RCTs and population-based assessments and attempted to determine the measures using published data from the trials.^17^ In addition, editorials by an editor associated with the BMJ also referred to this same lack of reporting.^18,19^ A few letters to editors of medical journals repeated the same points. During May 2020, after our formal literature review, one additional brief commentary appeared in a medical journal.^20^ In the general online media, a handful of critiques raised similar concerns.

### Risk reduction with vaccines

Table 1 shows data from the RCTs of Pfizer and Moderna and from the Israeli population-based study. The Pfizer and Moderna studies addressed the vaccines’ efficacy, which refers to their effects within controlled, experimental situations. The RRRs for both vaccines were in the range of 95 percent. This means that, among all those who became ill, 95 percent were in the unvaccinated group. The publications for these studies did not report ARRs, but data published in the studies permitted our calculation of ARRs. The calculated ARRs for vaccinated subjects compared to the baseline risk for unvaccinated subjects were in the range of 1 percent. The NNV to prevent one symptomatic infection, calculated from the ARR, was 141 for mild COVID and 2,500 for severe COVID in the Pfizer study, versus 88 and 500 in the Moderna study.

**Table 1.** Risk reduction with major vaccines.

|  |  | Control group<br>sample size (Nc) | Vaccinated group<br>sample size (Nv) |  |  |  |  |  |
| --- | --- | --- | --- | --- | --- | --- | --- | --- |
|  |  |  |  | Events in control<br>group (c) | Events in<br>vaccinated group<br>(v) | RRR=<br>[(c/Nc) - (v/Nv)]/<br>(c/Nc) | ARR=<br>(c/Nc) - (v/Nv) | NNV=1/ARR |
| A. Pfizer data |  |  |  |  |  |  |  |  |
|  |  | 21,720 | 21,728 |  |  |  |  |  |
|  | "Covid-19 with<br>onset at least 7 days<br>after the second<br>dose" |  |  | 162 | 8 | 95.06% | 0.71% | 141 |
|  | "severe Covid-19<br>with onset after the<br>first dose" |  |  | 9 | 1 | 88.89% | 0.04% | 2,500 |
| B. Moderna data |  |  |  |  |  |  |  |  |
|  |  | 15,210 | 15,210 |  |  |  |  |  |
|  | "symptomatic<br>COVID-19 illness" |  |  | 185 | 11 | 94.05% | 1.14% | 88 |
|  | "severe COVID" |  |  | 30 | 0 | 100.00% | 0.20% | 500 |
| C. Israel data |  |  |  |  |  |  |  |  |
|  |  | 596,618 | 596,618 |  |  |  |  |  |
|  | "documented<br>SARS-CoV-2<br>infection" |  |  | 6100 | 4460 | 26.89% | 0.27% | 370 |
|  | "symptomatic Covid-19" |  |  | 3607 | 2389 | 33.77% | 0.20% | 500 |
|  | "Covid-19 hospitalization" |  |  | 259 | 110 | 57.53% | 0.02% | 5,000 |
|  | "severe Covid-19" |  |  | 174 | 55 | 68.39% | 0.02% | 5,000 |
|  | "death due to Covid-19" |  |  | 32 | 9 | 71.88% | 0.004% | 25,000 |
| D. Smallpox, India data |  |  |  |  |  |  |  |  |
|  |  | 3,147 | 2,377 |  |  |  |  |  |
|  | Deaths |  |  | 944 | 76 | 89.34% | 26.80% | 4 |

Because the Israeli study assessed differences in symptomatic infections at the population level between vaccinated and unvaccinated persons, it measured the vaccines’ effectiveness in a “real-world” setting. RRRs were somewhat less impressive than in the RCTs (calculations based on Figure 2 in the published report): 34 percent for mild symptomatic disease, 58 percent for hospitalization, 68 percent for severe symptomatic disease, and 72 percent for death. The ARR from baseline risk was less than 1 percent. The NNV ranged from 500 for mild symptomatic infection to 25,000 for death.

### Measures of harm; harms versus benefits

Comparisons of harms versus benefits, as well as calculation of NNH, are challenging with currently available data. After two months of the RCTs, these studies permitted and encouraged vaccination among the control subjects, due to ethical concerns about withholding an apparently efficacious vaccine. For that reason, the usual longer follow up period to monitor for differences in the vaccines’ harms between subjects in the intervention and control groups did not occur. These studies did not report rates of harmful events consistently for vaccinated and control groups, and we were not able to ascertain denominators with numerical counts reported for some harmful events. The Pfizer study listed numerical counts for types of harm but provided comparative rates for vaccinated and control groups only for a few outcomes. In the Moderna study, the text presented numerical counts and percentages for some but not all adverse events. The Israel study did not present data on measures of harm.

Despite these limitations in available data on harm, we were able to calculate ARI and NNH for some data from the Pfizer and Moderna studies, as shown in Table 2. For minor adverse events, the ARIs in both studies were substantial, and the NNHs were less than 10. In the Pfizer study, the calculated ARI for a “serious adverse event” was 0.1%, and the NNH was 100. Considering “adverse events that were deemed by the trial team to be related to the vaccine or placebo,” the Moderna data showed a calculated ARI of 3.6% and an NNH of 27. For “treatment-related severe adverse events” in the Moderna study, the ARI was 0.3%, and the NNH was 333.

**Table 2.**
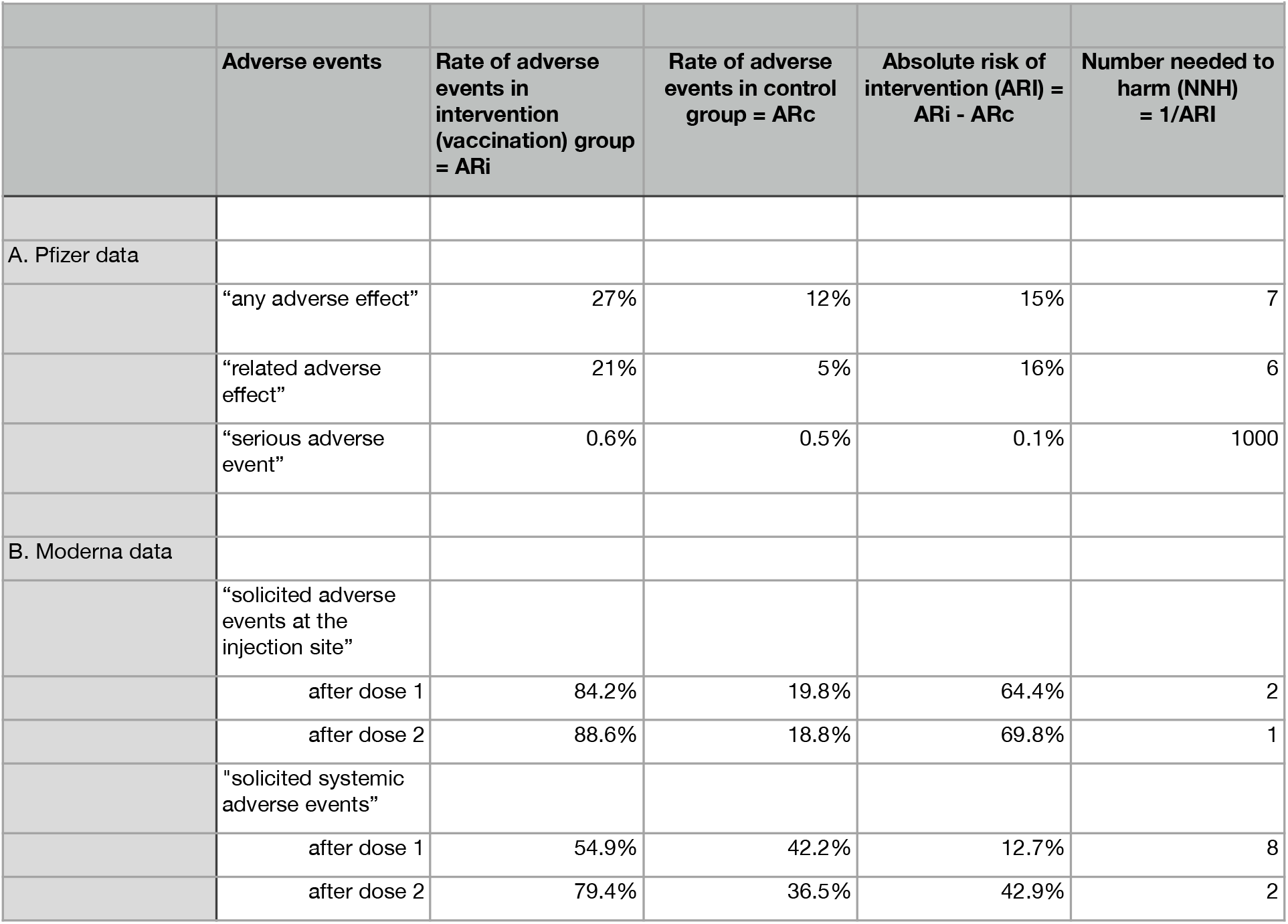
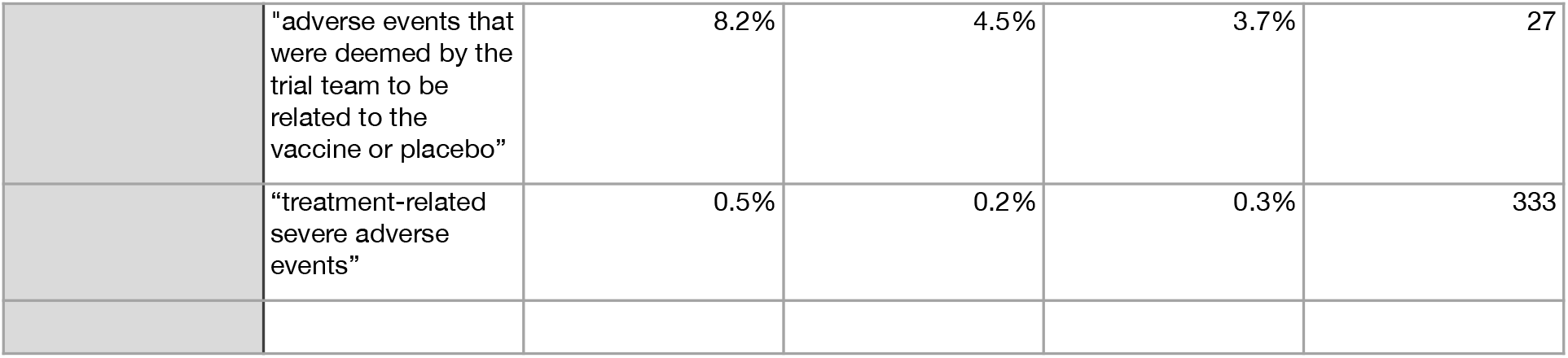
Measures of harm with major vaccine.

In this preliminary comparison of benefits and harms, the NNVs above from the three studies and the NNHs for “serious” or “severe” events from the Pfizer and Moderna studies appeared within a similar order of magnitude. Due to lack of longer term data on rates of harmful events from the three studies, we did not attempt to measure the significance of differences between NNV and NNH, for instance through confidence intervals. Other benefits from the vaccines may involve reduced transmission from vaccinated individuals to others and the possible development of herd immunity, but the magnitude of these effects attributable to the vaccines has not yet been determined.

### Vaccine benefits in populations with differing baseline risks

From the sensitivity analysis, the ARRs and NNVs are substantially more favorable in areas with high population baseline risk, as determined by cases per population (recognizing that this indicator usually is an underestimate based on incomplete case identification) (Table 3). Where the baseline risk is high, the ARR using a vaccine with high efficacy or effectiveness, as measured by RRR, is likely to be larger and the NNV smaller than where the baseline risk is low. For instance, if the baseline risk of infection in a region is 1.1250 percent (as shown for DeBaca County, NM, during May 2021) and about 50 percent of positives become symptomatic without vaccine (an assumption based on varying current estimates of this percentage^21,22^) the baseline risk of symptomatic disease is 0.5625 percent. Assuming vaccine efficacy as measured by RRR is 95 percent, the risk of symptomatic disease after vaccination is (0.05×0.5625 percent) = 0.0281 percent, the ARR is (0.5625-0.0281) = 0.5344 percent, and the NNV is (1/0.005344) = 187. But if the baseline risk of infection is 0.0080 percent (shown for Catron County, NM), the baseline risk of symptomatic disease is 0.0040 percent without vaccine and is reduced to 0.0002 percent with vaccine, for an ARR of 0.0038 percent and NNV of 2,601. Table 3 shows calculations for these and other selected U.S. counties and for states of India with differing baseline risks. In regions with lowest baseline risks, the NNV becomes much larger than in regions with highest baseline risks. For instance, in two states of India with high baseline risks, the NNV falls to the range of 30. In the Discussion section we discuss the policy implications of this analysis for priorities in vaccine distribution.

**Table 3.**
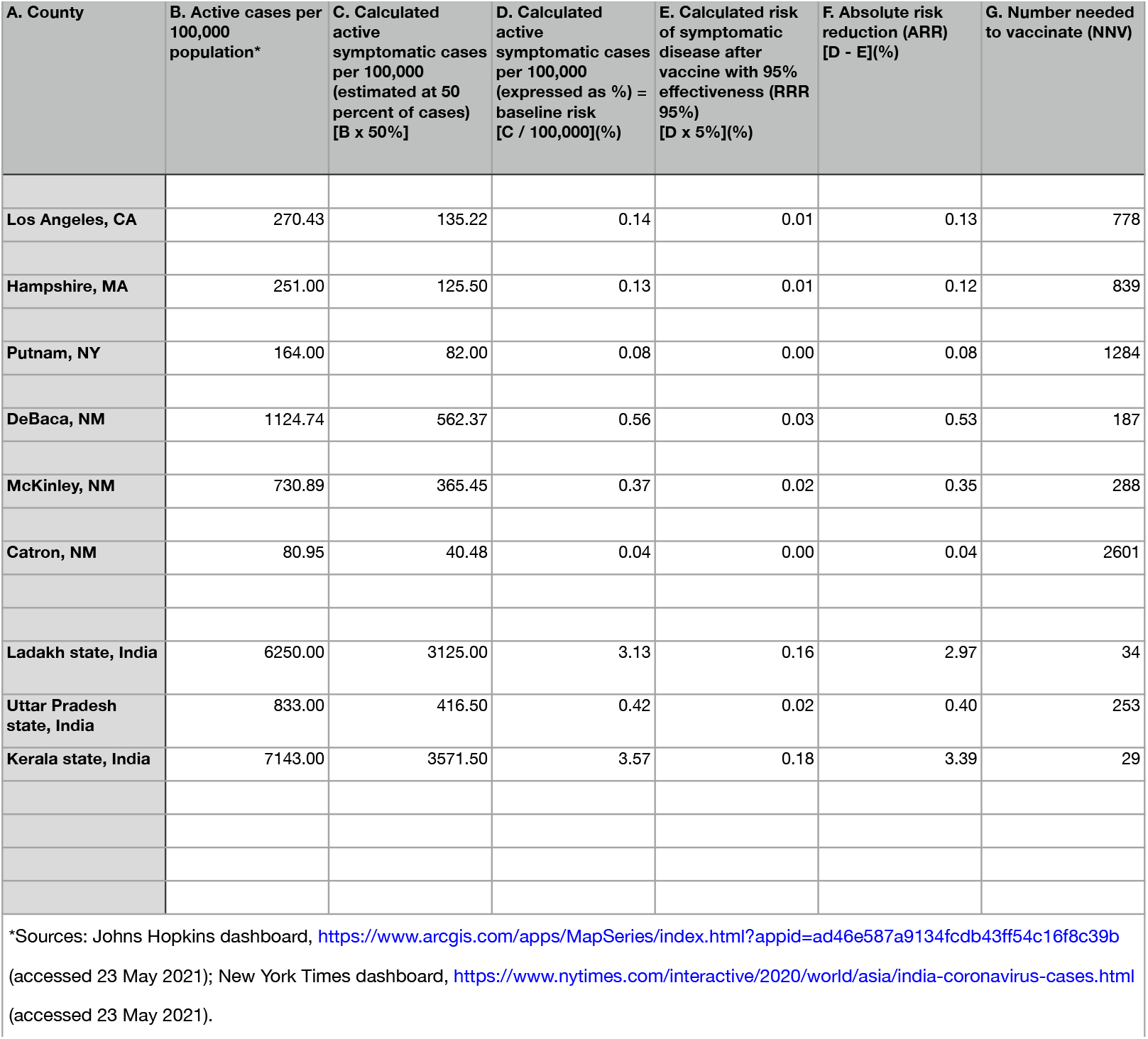
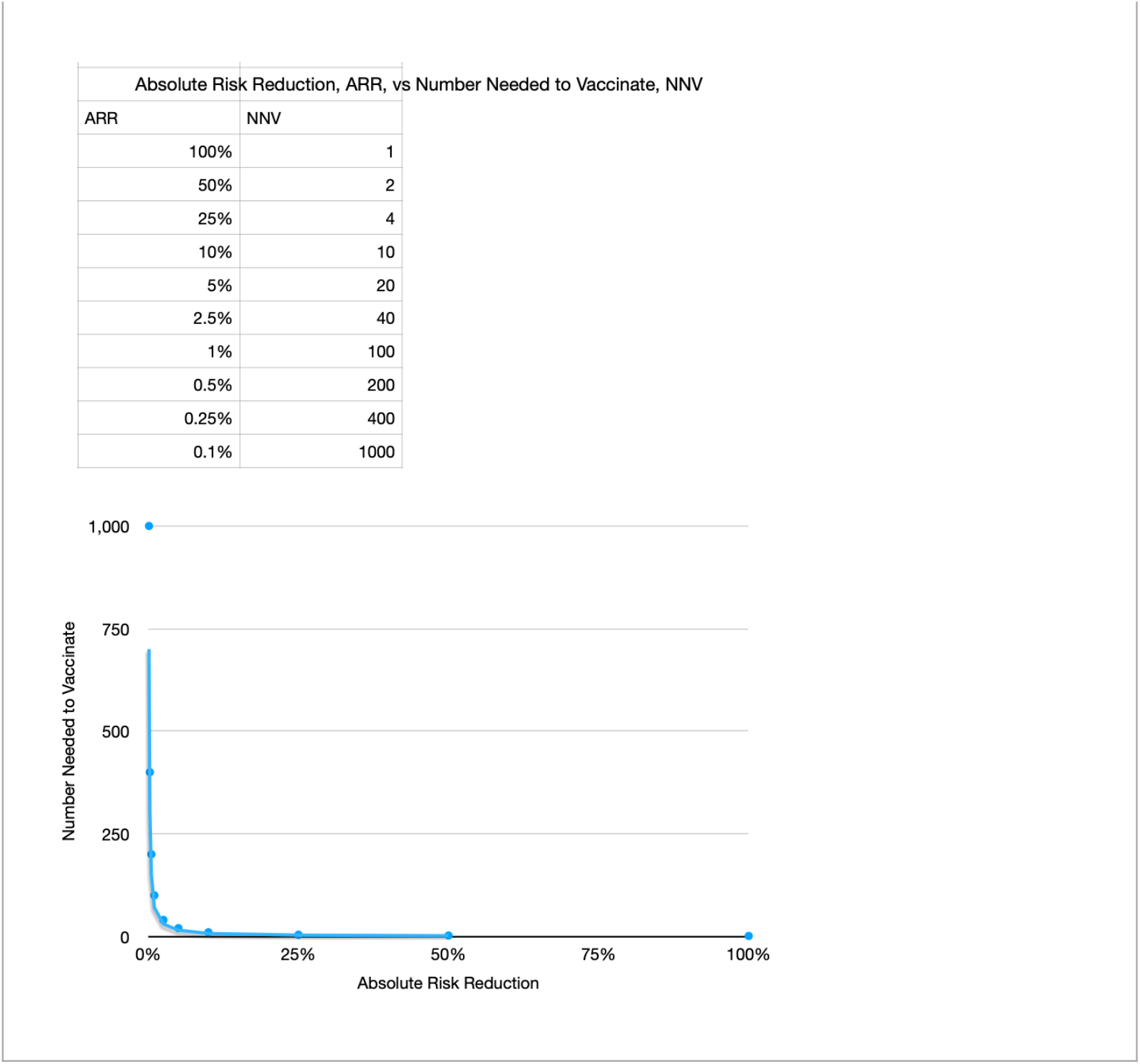
Numbers needed to vaccinate in geographical regions with different baseline risks; ARR versus NNV.

| A. County | B. Active cases per 100,000 population* | C. Calculated active symptomatic cases per 100,000 (estimated at 50 percent of cases) [B x 50%] | D. Calculated active symptomatic cases per 100,000 (expressed as %) = baseline risk [C / 100,000](%) | E. Calculated risk of symptomatic disease after vaccine with 95% effectiveness (RRR 95%) [D x 5%](%) | F. Absolute risk reduction (ARR) [D - E](%) | G. Number needed to vaccinate (NNV) |
| --- | --- | --- | --- | --- | --- | --- |
| Los Angeles, CA | 270.43 | 135.22 | 0.14 | 0.01 | 0.13 | 778 |
| Hampshire, MA | 251.00 | 125.50 | 0.13 | 0.01 | 0.12 | 839 |
| Putnam, NY | 164.00 | 82.00 | 0.08 | 0.00 | 0.08 | 1284 |
| DeBaca, NM | 1124.74 | 562.37 | 0.56 | 0.03 | 0.53 | 187 |
| McKinley, NM | 730.89 | 365.45 | 0.37 | 0.02 | 0.35 | 288 |
| Catron, NM | 80.95 | 40.48 | 0.04 | 0.00 | 0.04 | 2601 |
| Ladakh state, India | 6250.00 | 3125.00 | 3.13 | 0.16 | 2.97 | 34 |
| Uttar Pradesh state, India | 833.00 | 416.50 | 0.42 | 0.02 | 0.40 | 253 |
| Kerala state, India | 7143.00 | 3571.50 | 3.57 | 0.18 | 3.39 | 29 |

\*Sources: Johns Hopkins dashboard, https://www.arcgis.com/apps/MapSeries/index.html?appid=ad46e587a9134fdb43ff54c16f8c39b (accessed 23 May 2021); New York Times dashboard, https://www.nytimes.com/interactive/2020/world/asia/india-coronavirus-cases.html (accessed 23 May 2021).

| Absolute Risk Reduction, ARR, vs Number Needed to Vaccinate, NNV |  |
| --- | --- |
| ARR | NNV |
| 100% | 1 |
| 50% | 2 |
| 25% | 4 |
| 10% | 10 |
| 5% | 20 |
| 2.5% | 40 |
| 1% | 100 |
| 0.5% | 200 |
| 0.25% | 400 |
| 0.1% | 1000 |

### Comparisons with vaccines for other viral diseases

The ARRs and NNVs achieved by COVID vaccines so far appear somewhat less favorable than those achieved by vaccines for some other viral diseases. For instance, systematic reviews of influenza vaccines have revealed NNVs to prevent symptomatic infections between 12 and 94.^5^ Studies of herpes zoster vaccine have yielded NNVs for symptomatic infections between 11 and 43.^6^ For human papilloma virus vaccine, calculated NNV to prevent one case of cervical intraepithelial neoplasia in one study was 129.^23^ Regarding smallpox, the NNV to prevent one death was four (Table 1, calculated from data in ^24,25^). For such comparisons, limitations of NNV calculations include lack of consensus about targets for NNV across vaccines, questionable generalizability of findings from studies conducted in specific geographical regions, and inability to capture indirect benefits of vaccines such as their impact on transmission and development of herd immunity.^6^ Despite these limitations, current data indicate that available vaccines for COVID-19 may be reaching lower levels of effectiveness than some prior vaccines.

### How to explain these findings

Box 2 shows a brief explanation for possible use in informed consent procedures, patient education, continuing medical and public health education, and advice to policy makers.

## Discussion

### Research in context

From our systematic review, we found very few publications that refer to ARR and NNV in research on COVID-19 vaccines. In the medical literature, one article oriented to EBM and two editorials criticized the deficit in reporting of these measures in the major RCTs and population-based assessments. Although ARR and NNV have appeared in some recent publications about influenza vaccines, lack of attention to these key elements of EBM in assessments of medical interventions is not unique to assessments of COVID-19 vaccines. An overall review of reporting practices in the medical literature showed that among 875 controlled trials with binary outcomes and/or hazard ratios published in highly cited general medical journals, fewer than one-tenth reported at least one NNT or NNH, while slightly more than one-quarter reported at least one ARR.^26^ While the relative lack of attention to EBM measures in the medical literature raises concerns, this lack is especially troubling for trials of COVID-19 vaccines, due to the worldwide importance of clarity in assessing benefits such as efficacy and effectiveness, and also in assessing harms.

### Strengths and limitations of study

Our findings based on limited data available led to concerns about the vaccines’ impact in reducing absolute risk below the baseline risks for pertinent population groups, as well as about the comparative benefits versus harms of the vaccines. From our calculations using data from two major RCTs and one population-based assessment, we concluded that the impact of the vaccines in reducing the risk of symptomatic infection below the baseline risk as measured by ARR is small, and the NNV to prevent one symptomatic infection is substantial. Measures of harm and comparisons of harms versus benefits by calculation of ARI and NNH are challenging to achieve with published data from these trials. The NNHs that we calculated were in a range comparable to the benefits of the vaccines as indicated by NNVs. While we were not able to resolve the question of benefits versus harms, our limited analysis revealed that they were not dramatically different and that the NNHs were low for some outcomes. Compared to prior vaccines such as influenza and smallpox, the efficacy of the COVID-19 vaccines in the RCTs and effectiveness in the population-based study appeared somewhat less impressive. Importantly, the sensitivity analysis of the vaccines’ benefits showed that the ARRs and NNVs are more favorable in areas with higher baseline rates of disease. This variability in effectiveness related to varying baseline risks leads to some possibly helpful policy recommendations, as discussed further below. We were able to develop a preliminary template for explaining the findings to assist with obtaining informed consent, contributing to medical and public health educational efforts, and providing information for policy makers.

This study has several limitations. We have been able to analyze only the published data from the influential studies. It is not yet possible to access the raw data from these studies to determine if additional information might clarify with greater precision the ARRs and NNVs to assess vaccines’ benefits, as well as the ARIs and NNHs to assess harms. In particular, data on adverse effects from the vaccines were not available from one of the three studies considered. From the published reports, we could not determine if there was a rationale for not reporting ARR and NNV in addition to RRR, or for not reporting quantitative measures of harm such as NNH. The benefits of the vaccines include predictable effects in reducing transmission and fostering herd immunity, but those effects remain to be clarified through additional research. The degree to which the efficacy and effectiveness of the vaccines will change over time, especially as additional mutations arise and as public health practices such as social distancing and use of masks become different, remains unclear. Despite these limitations, our preliminary analysis shows that reductions of absolute risk and measures of harms versus benefits deserve more attention from the standpoint of EBM than they have received so far.

### Implications for research

We see several implications of this work for future research. First, in addition to RRR, research publications concerning the efficacy or effectiveness of vaccines should report other standard measures recommended in EBM, including ARR, NNV, ARI, and NNH. Investigators should try to compare benefits versus harms of vaccines, through measures like NNV and NNH. Based on ARR and NNV, the research reports should show how efficacy and effectiveness vary depending on baseline risks of disease in different population groups. The reports should provide practical advice for practitioners, patients, the general public, and policy makers that can guide informed consent procedures and decisions about vaccine distribution. Additional research also should try to illuminate the reasons why prominent publications about COVID-19 vaccines abandoned previously established reporting principles in EBM. Reasons for the exclusion by authors and editors of key measures like ARR and NNV warrant clarification. These reasons may include concerns about profitability of marketing vaccines, attempts to encourage hopeful attitudes about vaccination in the context of widespread desperation, and other concerns to be clarified, but the presence of any such motivations currently remains speculative.

### Implications for policy

Several implications for policy emerge from this work. Explanations to patients and the general public should present transparent information about ARR in addition to RRR. Informed consent procedures will remain incomplete unless educational strategies include such information. If a vaccine has effects on reducing transmission or enhancing herd immunity, these effects should be quantified and also explained clearly to patients and the general public. As shown by the sensitivity analysis of ARR and NNV, vaccine distribution initially should target geographic regions with higher baseline risks of disease, rather than focusing only on the goal vaccinating entire populations.

Assessments of baseline risks in small geographic areas, which rely on measures such as new cases per unit of time, overall active cases per population, and proportion of positive tests, are available from several sources, for instance, the Johns Hopkins University and *New York Times* dashboards.^15,27^ A strategy emphasizing vaccines’ differential impacts on reducing absolute risk in areas with varying baseline risks of disease could alleviate some economic and practical burdens of initially trying to provide vaccines for everyone, especially in poorer regions of richer countries, or in regions of poorer countries in the global South. While not in any way justifying unequal access to vaccines, this strategy is especially important as we face the reality of barriers to distribution related to wealth, power, minority status, structural racism, and other sources of inequality.

## Conclusion

In summary, some key principles of EBM have not guided the reports about research or approval of COVID-19 vaccines. These gaps have arisen especially in the quantification of impacts on absolute risk in studies of efficacy and effectiveness, which have emphasized high RRR; calculations of low ARR and substantial NNV provide important additional perspectives. Systematic comparisons of vaccines’ benefits versus harms have not emerged clearly from published reports, and preliminary analysis shows that benefits and harms are likely within a similar order of magnitude. Variations in vaccines’ impact on absolute risk depending on baseline risks of disease in different population groups should receive more attention in research and in policy recommendations about vaccine distribution. Such evidence-based principles gain even more importance in the context of barriers to vaccine access linked to profound socioeconomic inequality.

## Data Availability

All the data used in this study are publicly available and properly cited. More guided instruction to get access to the data for transparency and reproducibility will be provided on request.

## Contributors

AL conceptualized the study, with input from HW. AL and HW are the co-senior authors. AL coordinated the statistical analysis and systematic review, with inputs from HW. AL wrote the first draft of the manuscript, and HW did extensive revisions.

Both the co-authors approved of the final version of the manuscript. The corresponding author attests that all listed authors meet authorship criteria and that no others meeting the criteria have been omitted. HW is the guarantor.

## Ethical approval

As all the data were anonymous, and aggregated without any personal information, ethics approval was deemed waived.

## Data sharing

The authors affirm that the manuscript is an honest, accurate, and transparent account of the study being reported; that no important aspects of the study have been omitted; and that any discrepancies from the study as planned have been explained.

## Dissemination to participants and related patient and public communities

We will disseminate the findings widely to members of the public through official (press release, institutional websites, and repositories), personal, and social media. We also plan to write a op-ed and other opinion articles to describe this work in more general terms for the members of the public. The explanations developed for Box 2 will assist in these efforts.

## Provenance and peer review

Not commissioned; externally peer reviewed.

## Box 1.

**Definitions Used, Based on Evidence-Based Medicine**

- Absolute risk, AR, is the rate of an event (for instance, symptomatic COVID-19 infection) in a group.
- Absolute risk for the control group is designated ARc.
- Absolute risk for the intervention group is designated ARi.
- Absolute risk reduction: ARR is (ARc-ARi). This calculation shows the difference from a group’s baseline event rate between those receiving and not receiving an intervention; that is, it indicates the reduction of risk from the group’s baseline risk that results from the intervention. If c = events in control group and v = events in vaccinated group, Nc is the sample size of the control group, and Nv is the sample size of the vaccination group, ARR = (c/Nc) - (v/Nv).
- Relative risk reduction: RRR is (ARR/ARc) = (ARc-ARi)/ARc. This calculation shows the difference in event rate between those receiving and not receiving an intervention, expressed as a percentage of the event rate among those not receiving an intervention. If c = events in control group and v = events in vaccinated group, Nc is the sample size of the control group, and Nv is the sample size of the vaccination group, RRR = [(c/Nc) - (v/Nv)]/ (c/Nc).
- Number needed to vaccinate: NNV is 1/ARR. This calculation shows the number of people needed to be vaccinated in order to prevent a pertinent event in one person, such as a symptomatic infection by COVID-19.
- Absolute risk of an intervention: ARI is the rate of the intervention’s adverse effects, calculated by subtracting the rate of adverse effects in the group not receiving an intervention from that in the group receiving an intervention.
- Number needed to harm: NNH is 1/ARI. It shows how many people need to be vaccinated to cause harm for one person.
- Baseline risk is the total number of cases per population in a group not exposed to an intervention (such as vaccination) during a specified time period. In an RCT baseline risk is the proportion or percentage of study participants in the control group for whom a specified event, such as COVID-19 infection, is observed. In a community, baseline risk can be estimated by incidence (see below) among the unvaccinated population.
- Incidence is the number of new cases per population over a defined period of time.
- Prevalence is the total number of cases per population at one point in time.
- Efficacy is the degree to which a vaccination prevents an event under controlled conditions, as in an RCT.
- Effectiveness is the degree to which a vaccination prevents an event under “real-world” conditions, as in a population-based intervention.
- A risk-benefit assessment compares the risk of an intervention (for instance, NNH) to the benefit of an intervention (for instance, NNV).

## Box 2.

**Simplified Brief Explanation of Benefits and Harms of COVID 19 Vaccine**

### Patients and the general public

The available vaccines show a relative risk reduction (RRR) of about 50 percent to about 95 percent. This means that, if you get vaccinated with a vaccine showing highest RRR, your risk of getting COVID-19 symptoms will be about 5 percent of the risk for a person not getting the vaccine.

In making your decision, you also can consider some other important findings:

- The absolute risk of getting sick from COVID-19 depends on the baseline risk of COVID-19 infections in your population group. For instance, some people are members of groups with high rates of infection (people who work in meat packing plants, other “essential workers” who cannot work from home, and people who live in low-income and/or “minority” Black, Latinx, or Indigenous communities). If you are in a group with a high rate of infection, the absolute risk reduction (ARR) from getting the vaccine is greater than if you are in a group with a low rate of infection.

As an example, if the baseline risk of infection is 5 percent in your group, the likelihood that you’ll get sick with COVID-19 is 2.5 percent without a vaccine and 0.125 percent if you receive a vaccine that is 95 percent effective. So the vaccine will reduce your risk by 2.375 percent. If the baseline risk of infection is 2 percent in your group (which might apply if you are white, are well off, and can work at home), the likelihood that you’ll get sick with COVID-95 is 1 percent without a vaccine and 0.05 percent with a vaccine, a reduction of 0.95 percent. So the reduction in your risk by the vaccine will be greater if the baseline risk of infection is greater.

- From the studies so far, the likelihood of harm from the vaccine is real but small, less than 1 percent. If you are in a group with a higher baseline risk of COVID-19 infection, the benefit of the vaccine compared to the possible harm is larger than if you are in a group with a lower baseline risk.
- So we do recommend the vaccine for people in groups with higher baseline risks of disease. For groups with lower baseline risks, we are less sure that the benefit clearly outweighs the harm until we get more information over time.
- Some experts recommend the vaccine for everybody, partly based on the expected reduction of transmission and improved likelihood of herd immunity. But we actually don’t know yet how much these vaccines will reduce transmission, and we also don’t know much about the harms of the vaccines. Until we do know, we believe that individuals should make their decisions by considering how much a vaccine will reduce their risk below the baseline risk of disease for their population group.
- If you are in a population group with a high baseline risk of disease, we also recommend that you join with others in insisting that conditions decreasing your risk be addressed, such as masks and social distancing at work, and that services for communities – availability of adequate water supplies and soap for hand washing, adequate food, and housing that is not overcrowded – be improved.
- For everybody, you should not assume that receiving a vaccine removes the risk of getting COVID-19 or transmitting it to others. We recommend that everybody continue to apply basic public health principles to their own and others’ actions, especially about masks and maintaining adequate distancing to reduce risk of transmission.

### Health practitioners and policy makers

The most important message from our work is that the ARR from vaccines is largest in population groups with high baseline risks of COVID-19 infection. This understanding should shape priorities about vaccinations, both in clinical practice and in policy making. The inequalities in accessibility that have emerged worldwide mean that the vaccines will not reach large parts of the world’s population for the foreseeable future. Practitioners and policy makers must recognize that the benefits from the vaccines differ among different population groups and should prioritize those groups with high baseline risks of infection.

Especially in countries and regions where financial resources are insufficient to obtain and to deliver vaccines equally to entire populations, these resources initially should target those population groups with high baseline risks. This principle can help address the overwhelming problem of unequal resources among wealthy and less wealthy areas, both within and among countries. As more research findings become available over time about the relative benefits and harms of the vaccines, this information can guide subsequent decisions about the degree to which vaccines should be made available to population groups with lower baseline risks of disease.

## References

1 Laupacis A, Sackett D, Roberts R. An assessment of clinically useful measurements of the consequences of treatment. N Engl J Med 1988;318:1728–33.

2 Straus SE, Glasziou P, Richardson WS, Haynes RB. Evidence-Based Medicine: How to Practice and Teach EBM. Edinburgh: Elsevier 2019:90.

3 Fagerlin A, Peters E. Quantitative information. In: Fischhoff B, Brewer NT, Downs JS, eds. Communicating Risks and Benefits: An Evidence-Based User’s Guide. Washington DC: US Food and Drug Administration 2011:60. https://www.fda.gov/media/81597/download (accessed 23 Jun 2021).

4 Demicheli V, Jefferson T, Ferroni E, Rivetti A, Di Pietrantonj C. Vaccines for preventing influenza in healthy adults. Cochrane Database of Systematic Reviews 2018, Issue 2: Art. No.: CD001269. doi: 10.1002/14651858.CD001269.pub6.

5 Kolber MR, Lau D, Eurich D, Korownyk C. Effectiveness of the trivalent influenza vaccine. Can Fam Physician 2014;60:50. https://www.ncbi.nlm.nih.gov/pmc/articles/PMC3994812/pdf/0600050.pdf (accessed 23 Jun 2021).

6 Hashim A, Dang V, Bolotin S, Crowcroft NS. How and why researchers use the number needed to vaccinate to inform decision making—A systematic review. Vaccine. 2015;33:753–8. http://dx.doi.org/10.1016/j.vaccine.2014.12.033.

7 Page MJ, McKenzie JE, Bossuyt PM, et al. The PRISMA 2020 statement: an updated guideline for reporting systematic reviews. BMJ 2021;372:71 http://dx.doi.org/10.1136/bmj.n71.

8 JAMA evidence. Glossary. https://jamaevidence.mhmedical.com/DocumentLibrary/JAMAevidence_Glossary_Final.pdf (accessed 23 Jun 2021).

9 University of Oxford, Center for Evidence-Based Medicine. Number needed to treat (NNT). https://www.cebm.ox.ac.uk/resources/ebm-tools/number-needed-to-treat-nnt (accessed 23 Jun 2021).

10 Statistics How To. Relative risk and absolute risk: definition and examples. https://www.statisticshowto.com/calculate-relative-risk/ (accessed 23 Jun 2021).

11 Arlegui H, Bollaerts K, Salvo F, et al. Benefit–risk assessment of vaccines. Part I: A systematic review to identify and describe studies about quantitative benefit–risk models applied to vaccines. Drug Safety 2020;43:1089–1104. https://doi.org/10.1007/s40264-020-00984-7.

12 Polack FP, Thomas SJ, Kitchin N, et al. Safety and efficacy of the BNT162b2 mRNA Covid-19 vaccine. N Engl J Med 2020;383:2603–15. doi: 10.1056/NEJMoa2034577.

13 Baden LR, El Sahly HM, Essink B, et al. Efficacy and safety of the mRNA-1273 SARS-CoV-2 vaccine. N Engl J Med 2021;384:403–41. doi: 10.1056/NEJMoa2035389.

14 Dagan N, Barda N, Kepten E, et al. BNT162b2 mRNA Covid-19 vaccine in a nationwide mass vaccination setting. N Engl J Med 2021;384:1412–23. doi: 10.1056/NEJMoa2101765.

15 COVID-19 trends for U.S. counties. Johns Hopkins University COVID-19 dashboard. https://www.arcgis.com/apps/MapSeries/index.html?appid=ad46e587a9134fcdb43ff54c16f8c39b (accessed 25 Jun 2021).

16 Maps and cases. The coronavirus outbreak. New York Times. https://www.nytimes.com/interactive/2021/us/covid-cases.html?name=styln-coronavirus&region=TOP_BANNER&block=storyline_menu_recirc&action=click&pgtype=LegacyCollection&variant=1_Show&is_new=false (accessed 25 June 2021).

17 Brown RB. Outcome reporting bias in COVID-19 mRNA vaccine clinical trials. Medicina 2021;57:199. https://doi.org/10.3390/medicina5703 0199.

18 Doshi P. Pfizer and Moderna!s “95% effective” vaccines—let!s be cautious and first see the full data. BMJ 2020 (Nov 26). https://blogs.bmj.com/bmj/2020/11/26/peter-doshi-pfizer-and-modernas-95-effective-vaccines-lets-be-cautious-and-first-see-the-full-data/ (accessed 23 Jun 2021).

19 Doshi P. Pfizer and Moderna!s “95% effective” vaccines—we need more details and the raw data. BMJ 2021 (Jan 4). https://blogs.bmj.com/bmj/2021/01/04/peter-doshi-pfizer-and-modernas-95-effective-vaccines-we-need-more-details-and-the-raw-data/ (accessed 23 Jun 2021).

20 Olliaro P, Torreele E, Vaillant M. COVID-19 vaccine efficacy and effectiveness—the elephant (not) in the room. Lancet Microbe 2021 (April 20). https://doi.org/10.1016/S2666-5247(21)00069-0.

21 Oran DP, Topol EJ. Prevalence of asymptomatic SARS-CoV-2 infection: A narrative review. Ann Intern Med 2020;173:362–7. doi: 10.7326/M20-3012.

22 Yanes-Lane M, Winters N, Fregonese F, et al. Proportion of asymptomatic infection among COVID-19 positive persons and their transmission potential: A systematic review and meta-analysis. PLoS ONE 15(11): e0241536. https://doi.org/10.1371/journal.pone.0241536.

23 Sawaya GF, Smith-McCune K. HPV vaccination – more answers, more questions. N Engl J Med. 2007;356:1991–3. doi: 10.1056/NEJMe078060.

24 Petersen BW, Damon IK, Pertowski CA, et al. Clinical guidance for smallpox vaccine use in a postevent vaccination program. MMWR Recomm Rep 2015;64(RR02):1–26. https://www.cdc.gov/mmwr/pdf/rr/rr6402.pdf (accessed 23 Jun 2021).

25 Conti R. Smallpox vaccinations: The risks and the benefits. Issue Brief (Commonw Fund) 2003;620:1–9. [PMID: 12693396]. https://www.ncbi.nlm.nih.gov/pmc/articles/PMC4598063/ (accessed 23 Jun 2021).

26 Elliott MH, Skydel JJ, Dhruva SS, Ross JS, Wallach JD. Characteristics and reporting of number needed to treat, number needed to harm, and absolute risk reduction in controlled clinical trials, 2001-2019. JAMA Intern Med 2021;181:282–4. doi: 10.1001/jamainternmed.2020.4799.

27 Lutz E, Aufrichtig E, Smart C, Sun A, Harris F, Gianordoli G. A guide for Covid-19 risk in your county. New York Times Dashboard. https://www.nytimes.com/interactive/2021/us/covid-risk-map.html#methodology (accessed Jun 23, 2021).

